# Epidemiological Profile of Road Traffic Injury Patients in Hospitals in Fako Division, Cameroon: A Foundation for Tailored Post-Traumatic Stress Disorder (PTSD) Management

**DOI:** 10.1101/2025.06.10.25329319

**Authors:** Claudia Ngeha Ngu, Priya Shete, Dickson Shey Nsagha, Elvis Asangbeng Tanue, Nicholas Tendongfor, Rasheedat Oke, Nahyeni Bassah, Chrisantus Eweh Ukah, Sandra I. McCoy, Catherine Juillard, Alain Chichom Mefire, Edie Gregory Halle Ekane

**Affiliations:** Department of Public Health and Hygiene, Faculty of Health Sciences, University of Buea, Buea, Cameroon; Department of Medicine, University of California San Francisco, San Francisco, CA, USA; Program for the Advancement of Surgical Equity, Department of Surgery, University of California Los Angeles, Los Angeles, California, USA; Division of Epidemiology, School of Public Health, University of California Berkeley, California, USA

**Keywords:** Road Traffic Injury, Epidemiological Profile, Cameroon, Post Traumatic Stress Disorder, Fako, Mental Health

## Abstract

**Background:** Road traffic injuries (RTIs) are a major global public health concern, often leading to serious physical and psychological consequences, including post-traumatic stress disorder (PTSD). A limited understanding of specific RTI patient characteristics particularly regarding PTSD screening recommendation and uptake in Cameroon hinders, the development of effective PTSD management strategies. This study aimed to determine the epidemiological profile of RTI patients in Fako Division, Cameroon, to inform the development of tailored PTSD management interventions.

**Material and methods:** A chart review of medical records of RTI patients received between 2019 and 2023 was conducted from July 30^th^ 2024 to August 30^th^ 2024 at three hospitals in Fako Division, Cameroon: Buea Regional Hospital, Saint Luke Hospital Buea, and Limbe Regional Hospital. A data extraction form was used to collect demographic information, injury characteristics, and PTSD screening recommendations and uptake. Descriptive statistics and Chi square test were used for data analysis. The data was analysed in SPSS version 25.

**Results:** A total of 4218 RTI patients were received and managed at the three hospitals during the study period. Patients were predominantly male, 2937(69.6%). The age group of 26-35 years recorded the highest proportion, 1339(31.7%). The median age of patients was 28 years (interquartile range: 22-37). Students, 820(19.4%) were among the most involved in RTIs. Only 59(11.0%) out of the 534(12.7%) hospitalized patients after RTIs were recommended for PTSD screening, and only 26(44.1%) underwent PTSD screening. Lower limbs, 2136(50.6%), and upper limbs, 1639(38.9%) were the most affected location for the traumatic injury. In-hospital mortality was recorded in 27(5.1%) of the patients. Means of transportation of patient to the hospital, (*p*=0.050), pain medication given (*p*=0.024), sedative given (*p*= **<**0.001), surgery performed (*p*=0.005), psychotherapy recommended (*p*=**<**0.001) and transfusion performed (*p*=<0.001) were significantly associated with recommendation for PTSD screening. A statistically significant association was found between PTSD screening uptake and recommendation for psychotherapy (*p*=0.010).

**Conclusion:** Few patients were recommended for PTSD screening and uptake was low. This indicates a significant gap in access to PTSD and mental health services for RTI patients in Fako. PTSD screening uptake was significantly associated with recommendation for psychotherapy. Findings provide information for developing tailored PTSD management strategies and improving patients’ outcomes in this region.

## Introduction

Road traffic injuries (RTIs) represent a major global public health challenge, causing over 1.19 million deaths and around 50 million injuries each year, with a particularly heavy toll on low- and middle-income countries (LMICs) due to their higher rates of long-term disability, morbidity, and associated economic burdens [1, 2]. This number is projected to rise to two million deaths per year by 2030 [3]. In Cameroon, a LMIC, the situation of RTI is dire. The number of RTI is on the rise and the burden is reflected in the estimated 1443 disability-adjusted life years (DALYs) per 100,000 population lost to road crashes [4]. From Government statistics, Cameroon records an estimate of 16,583 road crashes each year, killing over 1,000 people, and approximately 6000 according to WHO estimates [10-12]. A pilot study in Cameroon reported that injuries constitute a significant proportion of emergency visits and utilization of surgical services [7]. Sociopolitical and economic reasons have also led to an increased presence of commercial motorcycles over the years in Cameroon as a whole and Fako Division in particular. This has exacerbated the situation as motorcycles are mostly involved in RTIs [6, 7]. Cameroon, characterized by rapid urbanization, limited enforcement of road safety regulations, and availability of motorcycles as an affordable and accessible means of transport has coincided with this increase in road traffic crashes [17, 18].

In addition to the physical injuries to victims, RTI can have serious psychological consequences not just for the RTI victims but for their families as well [3, 4]. A significant proportion of RTI survivors develop psychological disorders after a road crash such as post-traumatic stress disorder (PTSD), major depressive disorder, phobia for driving and/ or traveling, and other anxiety disorders [5, 6]. Of these, PTSD is a prevalent psychological disorder following RTIs [7, 8]. The pooled prevalence of PTSD among RTI victims globally is 22.2%, and studies show that 1 in 4 RTI victims in Africa develop PTSD [15, 17]. Previous studies show that survivors do not recover to their pre-accident state for several years after RTI [9-11]. In Cameroon, mental health has been integrated into the minimum package of healthcare services in line with WHO recommendations [20]. Studies show that Psychiatrists are predominantly clustered in major cities like Douala and Yaounde in the Littoral and Center regions respectively [20, 21], highlighting concerns about mental health care provision in other regions, such as the South West Region and Fako division in particular. Cameroon faces unique challenges in mitigating the impact of RTIs and providing adequate care for affected individuals, this is further compounded by the scarcity of mental health services [22].

Despite the substantial burden of RTIs in Cameroon, and the high prevalence of PTSD after RTI, data on RTIs in Cameroon remain sparse resulting in a limited understanding of specific RTI patient characteristics, particularly regarding PTSD screening recommendations and uptake in Cameroon. Within Cameroon, data on the epidemiological characteristics of RTI patients, and associated mental health conditions such as PTSD remain scarce, especially at the sub-national level with previous research focused on describing RTI patients’ demographic and injury characteristics [21-23]. More so, though mental health services have been integrated into primary healthcare, there is a dearth of information on the effective utilization of these services with regard to PTSD screening recommendation and uptake among RTI victims. To our knowledge, no study in this setting has reviewed RTI data on psychological care such as PTSD in RTI. This gap in knowledge hinders the development of effective evidence-based PTSD management strategies and interventions. More so, attention of care by health care providers is on physical injury with less attention given to the psychological consequences experienced by RTI survivors, with a frequent oversight of Post-Traumatic Stress Disorder, a prevalent mental health condition after RTI. This gap in mental health care can severely compromise the recovery and overall well-being of victims [25]. This oversight is particularly concerning in LMICs like Cameroon where limited resources and competing health priorities often overshadow mental health needs. Furthermore, the epidemiological patterns and psychological sequelae of RTIs can vary across different populations. Factors such as socioeconomic conditions, road infrastructure, access to emergency medical care, and cultural beliefs about mental health all influence the experience of RTI survivors and the likelihood of developing PTSD [26]. There is therefore a growing need to understand RTI survivors’ specific epidemiological profile and factors that are associated with recommendations for PTSD screening and PTSD screening uptake.

This gap in knowledge underscores the need for targeted research to better understand the dynamics of RTIs and mental health sequelae such as PTSD within the Cameroonian context. As such, the Fako Division represents an essential setting for this research to better inform PTSD management in the region.

The main aim of this study was to describe the epidemiological profile of road traffic injury patients in hospitals in Fako Division-Cameroon, 2019-2023, particularly with regard to PTSD screening recommendation and uptake. This data will be used to inform tailored Post-Traumatic Stress Disorder management strategies. Understanding the demographic and clinical profiles of RTI patients is important in identifying factors associated with PTSD screening recommendation and uptake from patient profiles and identifying gaps in PTSD management which can guide early detection and intervention. More so, it will provide data to advocate for integrated trauma-mental health services based on local evidence.

## Methods

### Study design, setting, and data source

This was an exhaustive chart review of medical records of RTI patients received and managed between January 2019 and December 2023 at three purposefully selected hospitals in Fako Division, Cameroon: Buea Regional Hospital, Saint Luke Hospital Buea, and the Limbe Regional Hospital. The chart review was done between July 30^th^ 2024 to August 30^th^ 2024. These hospitals are either trauma centers or noted for receiving many RTI victims. Fako Division is densely populated due to factors like the presence of; universities, touristic sites, oil refinery, and plantations, leading to an increase presence of vehicles, and increased RTI risk as the road infrastructure is not the best. The Limbe Regional Hospital is a 200-bed trauma center and referral hospital with an emergency and casualty department, an outpatient department, and a general surgery unit. Saint Luke Hospital offers orthopedic services to trauma patients including RTI victims. Buea Regional Hospital is a 120-bed capacity referral hospital with an orthopedic department, surgical unit, emergency unit, outpatient department, and mental health unit for trauma care [11, 12].

### Data collection

Trained data collectors systematically identified and reviewed hospital records (consultation, hospitalization, and trauma registers, as well as RTI patients’ files) from relevant units involve with the care of RTI victims, including emergency, surgery, intensive care, orthopedics, and outpatient departments. We included records of all RTI patients received and/ or managed in these three health facilities from January 2019 to December 2023. A data extraction form designed using Kobo toolbox software was used. This form was pretested at the Solidarity Clinic, a hospital in Buea with experience in managing RTI cases, allowing us to refine the tool and ensure suitability for our study. Records of patients that met our inclusion criteria in these hospitals were reviewed and data was exhaustively extracted and entered into the kobo toolbox software. Incomplete files were excluded. Demographic characteristics, injury-related characteristics, and hospital care characteristics including PTSD screening recommendation and PTSD screening uptake data were extracted.

### Ethical considerations

Ethical approval for the study was obtained from the Institutional Review Board of the Faculty of Health Sciences of the University of Buea (No: 2024/2524-04/UB/SG/IRB/FHS). To maintain confidentiality since authors had access to information that could identify individual participants during data collection, a code was assigned to each reviewed RTI patient’s record for de-identification.

### Data analysis

Data were retrieved from Kobo Toolbox, exported to Microsoft Excel version 13 for initial cleaning, and subsequently imported into SPSS version 25 for statistical analysis. Categorical variables were presented as frequencies and percentages. Continuous variables were presented as means (and standard deviation). Descriptive statistics were generated, resulting in an epidemiological profile of RTI patients based on: demographics, injury, and hospital care characteristics. Chi-square test or Fisher’s exact test (when at least an expected count in a cell was less than 5) was applied to identify associations between demographic characteristics, injury characteristics, hospital care characteristics, and PTSD screening recommendation and uptake. A *p*-value of less than 0.05 was considered statistically significant. Results were represented in tables and figures.

## Results

### Socio-demographic characteristics of participants

A total of 4,218 RTI patients were received and managed at the three hospitals during the study period of January 2019-December 2023. Majority, 3,854 (91.4%), were received at the Limbe Regional hospital. Patients were predominantly male 2,937 (69.6%). The age group, 26-35 years recorded the highest proportion of cases, 1339 (31.7%). The mean age of patients was 30.34 (±13.51) years. Students, 820 (19.4%) were among the most involved (Table 1).

**Table 1:** Socio-demographic Characteristics of RTI Patients, Fako Division, 2019-2023 (n= 4218)

| Variable | Category | Frequency (%) |
| --- | --- | --- |
| Hospital | Buea Regional Hospital | 321 (7.6) |
|  | Saint Luke Hospital | 43 (1.0) |
|  | Limbe Regional Hospital | 3854 (91.4) |
| Age (years) | <16 | 427 (10.1) |
|  | 16-25 | 1226 (29.1) |
|  | 26-35 | 1339 (31.7) |
|  | 36-45 | 686 (16.3) |
|  | 46-55 | 316 (7.5) |
|  | 56-65 | 158 (3.7) |
|  | >65 | 66 (1.6) |
| Sex | Female | 1281 (30.4) |
|  | Male | 2937 (69.6) |
| Employment Status | Employed | 2792 (66.2) |
|  | Retired | 67 (1.6) |
|  | Student | 820 (19.4) |
|  | Unemployed | 539 (12.8) |

### Injury Characteristics of Study Participants

The crashes frequently occurred at night, 356 (41.7%). Of the 928 records capturing types of vehicles involved, motorcycles comprised more than half, 498(53.7%). The highest number of RTIs was recorded in 2022, 975 (23.1%). The most used means of transportation for RTI patients to the hospital was 2-3 wheeled vehicles, 1451(56.7%), and ambulance constituted only 42 (1.6%) (Table 2). Lower limbs, 2136 (50.6%), and upper limbs, 1639 (38.9%) were the most affected anatomical locations for the traumatic injury (Figure 1).

**Figure 1:**
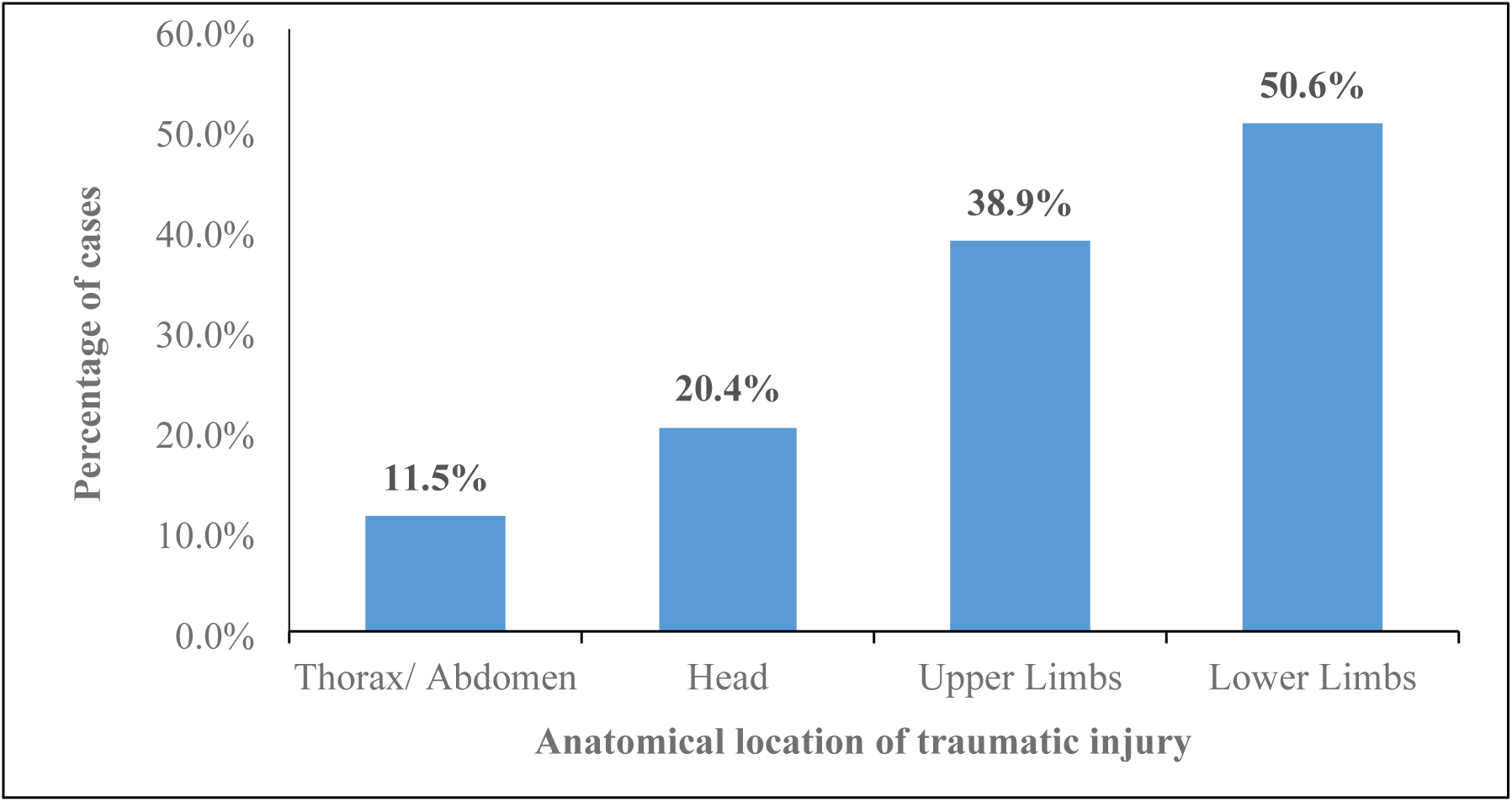
Distribution of RTIs by anatomical location of traumatic injury, Fako, 2019-2023.

**Table 2:**
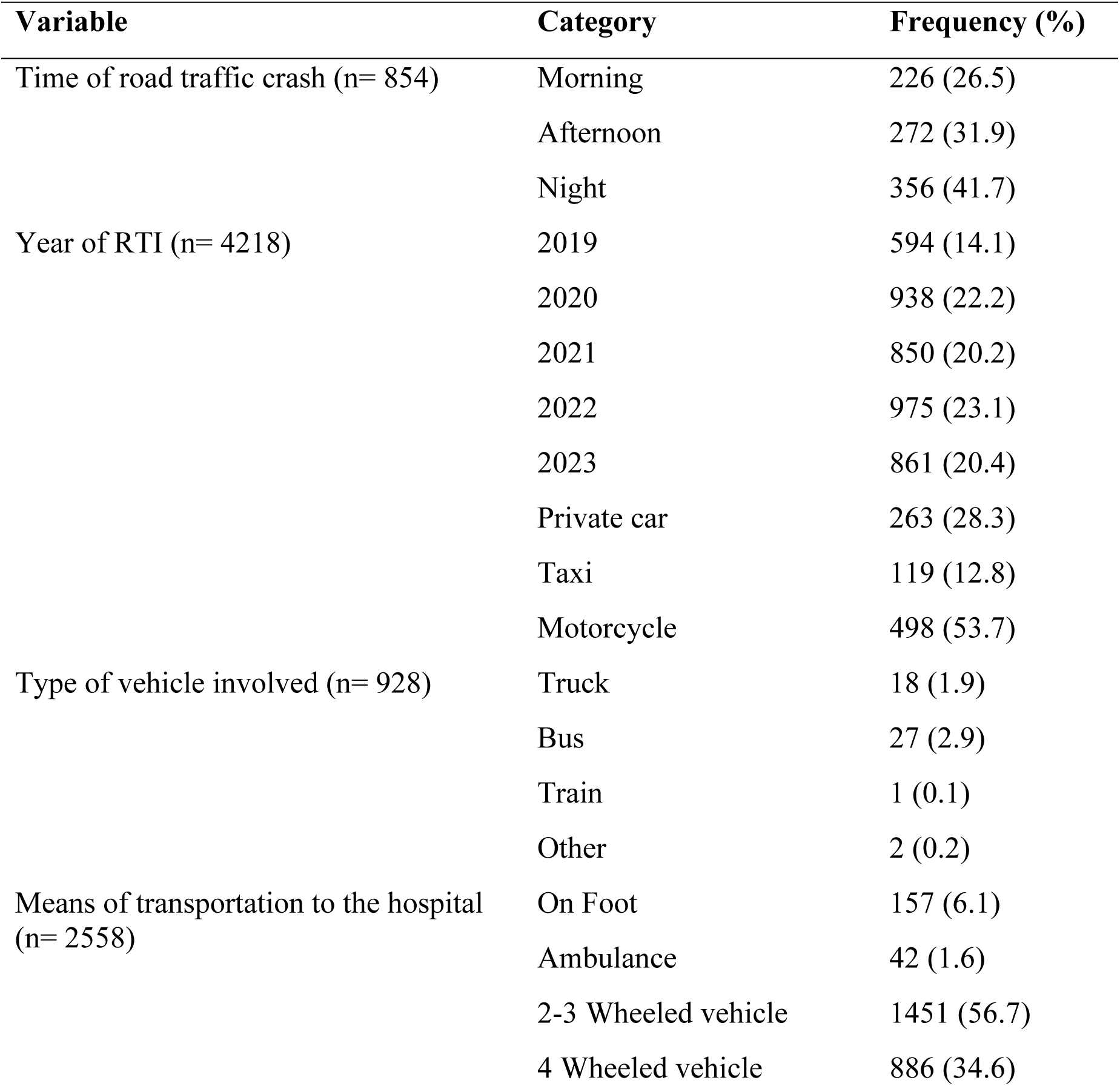
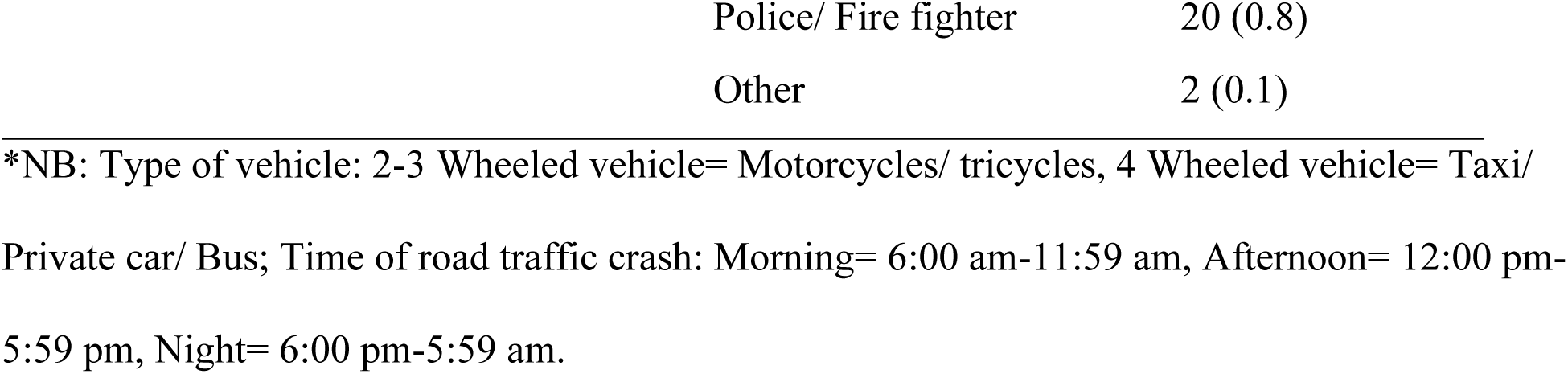
Injury Characteristics of RTI Patients in the Fako Division, 2019-2023.

### In-Hospital Care Received and Outcomes of Road Traffic Injury Patients

Five hundred and thirty-four (12.7%) RTI patients were hospitalized, and 76 (1.7%) were referred. Of the admitted cases, 500 (93.6%) were given pain medication, and 348 (65.2%) administered sedative. About half of the hospitalized patients, 280 (52.4%) underwent surgery. Only 59 (11.0%) of hospitalized patients were recommended for PTSD screening. PTSD screening uptake was only 26 (44.1%) among those recommended for PTSD screening. Very few patients were recommended for psychotherapy, 47 (8.8%) and the uptake was 40 (85.1%) among those recommended. Two hundred and three (38.0%) patients received transfusion. Regarding patients’ outcomes; In-hospital mortality was recorded in 27(5.1%) of the patients, 456(85.4%) were discharged home, 21(3.9%) were referred/ transferred to other hospitals, and 30(5.6%) patients were discharged against medical advice (Table 3).

**Table 3:**
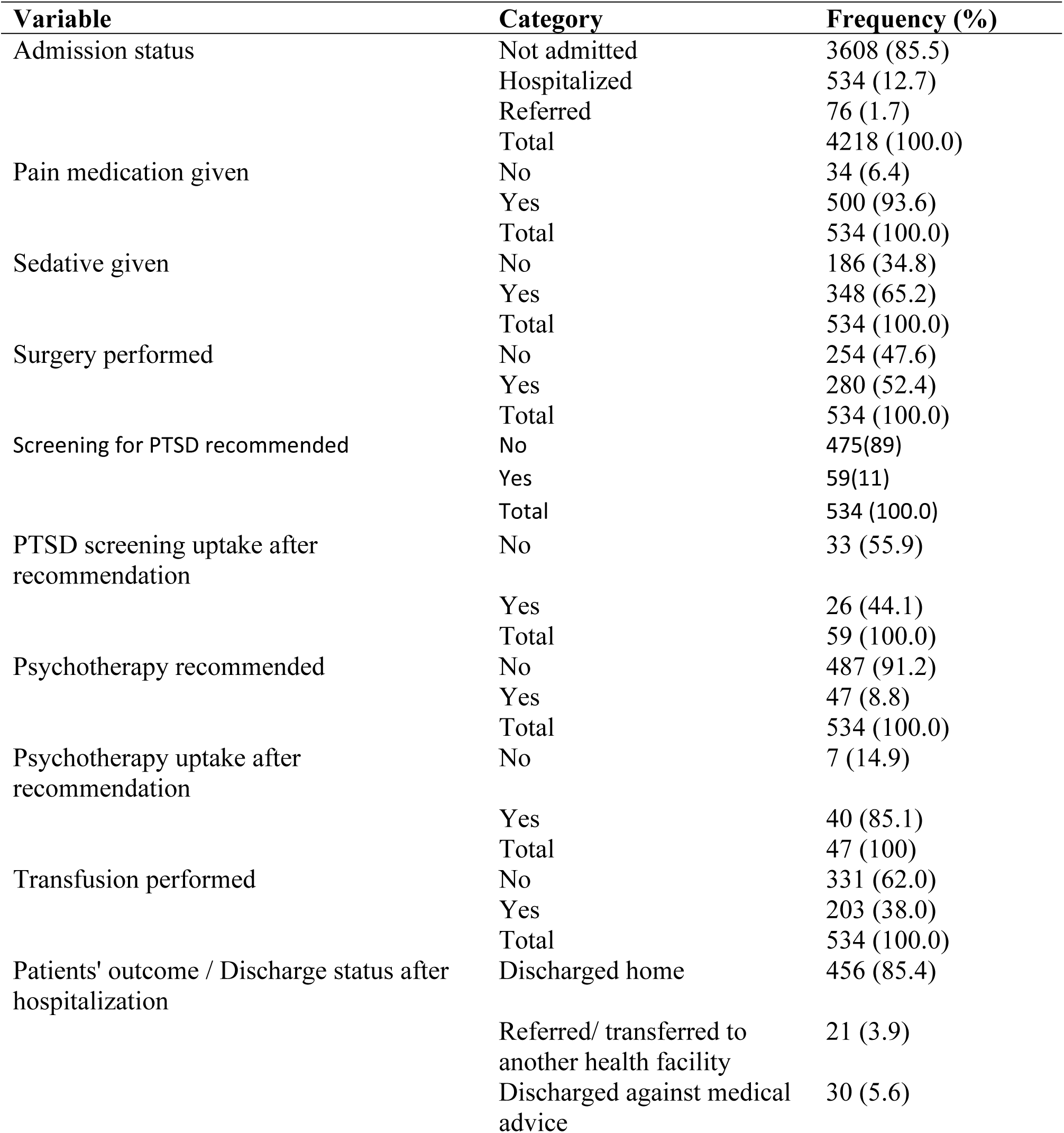
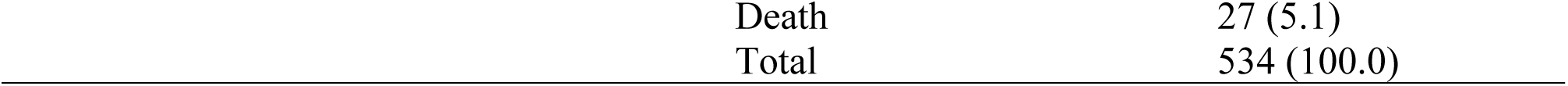
In-Hospital Care received and Outcomes of RTI patients, Fako, 2019-2023.

### Association between Recommendation for PTSD Screening and RTI Characteristics

On bivariate analysis using Chi square test; means of transportation of the patient to the hospital, (*p*= 0.050), pain medication given (*p*= 0.024), sedative given (*p*= **<**0.001), surgery performed (*p*= 0.005), psychotherapy recommended (*p*= **<**0.001) and transfusion performed (*p*= <0.001) were significantly associated with recommendation for PTSD screening. Of the 59 (11.0%) hospitalized RTI patients who were recommended for PTSD screening, 53(89.8%) were discharged home. No statistically significant association was found between recommendation for PTSD screening and RTI patients’ outcome (*p*= 0.902) (Table 4).

**Table 4:** Association between Recommendation for PTSD Screening and RTI Characteristics.

| Variable | Category | PTSD screening recommended |  | Total | Chi Square | P-Value |
| --- | --- | --- | --- | --- | --- | --- |
|  |  | Frequency (%) |  |  |  |  |
|  |  | No | Yes |  |  |  |
| Means of transport to the hospital | On Foot | 4(1.7) | 4(6.9) | 8(2.7) | 9.619 | **0.050\*** |
|  | Ambulance | 15(6.4) | 5(8.6) | 20(6.8) |  |  |
|  | 2-3 Wheeled vehicle | 92(39.1) | 15(25.9) | 107(36.5) |  |  |
|  | 4 Wheeled vehicle | 117(49.8) | 34(58.6) | 151(51.5) |  |  |
|  | Police/ Fire fighter | 7(3.0) | 0(0.0) | 7(2.4) |  |  |
|  | Total | 235(100) | 58(100) | 293(100) |  |  |
| Pain medication given | No | 34(7.2) | 0(0.0) | 34(6.4) | 4.510 | **0.024\*** |
|  | Yes | 441(92.8) | 59(100) | 500(93.6) |  |  |
|  | Total | 475(100) | 59(100) | 534(100) |  |  |
|  |  | Frequency (%) |  |  |  |  |
|  |  | No | Yes |  |  |  |
| Sedative given | No | 184(38.7) | 2(3.4) | 186(34.8) | 28.887 | <0.001 |
|  | Yes | 291(61.3) | 57(96.6) | 348(65.2) |  |  |
|  | Total | 475(100) | 59(100) | 534(100) |  |  |
| Surgery performed | No | 236(49.7) | 18(30.5) | 254(47.6) | 7.737 | 0.005 |
|  | Yes | 239(50.3) | 41(69.5) | 280(52.4) |  |  |
|  | Total | 475(100) | 59(100) | 534(100) |  |  |
| Psychotherapy recommended | No | 460(96.8) | 27(45.8) | 487(91.2) | 170.589 | <0.001 |
|  | Yes | 15(3.2) | 32(54.2) | 47(8.8) |  |  |
|  | Total | 475(100) | 59(100) | 534(100) |  |  |
| Transfusion performed | No | 320(67.4) | 11(18.6) | 331(62.0) | 52.876 | <0.001 |
|  | Yes | 155(32.6) | 48(81.4) | 203(38) |  |  |
|  | Total | 475(100) | 59(100) | 534(100) |  |  |
| Discharge status | Discharged home | 403(84.8) | 53(89.8) | 456(85.4) | 1.153 | 0.902* |
|  | Referred | 19(4.0) | 2(3.4) | 21(3.9) |  |  |
|  | Discharged against medical advice | 28(5.9) | 2(3.4) | 30(5.6) |  |  |
|  | Death | 25(5.3) | 2(3.4) | 27(5.1) |  |  |
|  | Total | 475(100) | 59(100) | 534(100) |  |  |

NB:\*= Fisher's Exact Test, 2-3 wheeled vehicle= motorcycles/bicycles/tricycles, 4 Wheeled vehicle =Taxi/Private car/Bus.

### Association between PTSD Screening Uptake after Recommendation, and RTI Patients’ characteristics

A statistically significant association was found between PTSD screening uptake and recommendation for psychotherapy (*p*= 0.010) with 19 (73.1%) patients who were screened for PTSD recommended for psychotherapy (Table 5).

**Table 5:**
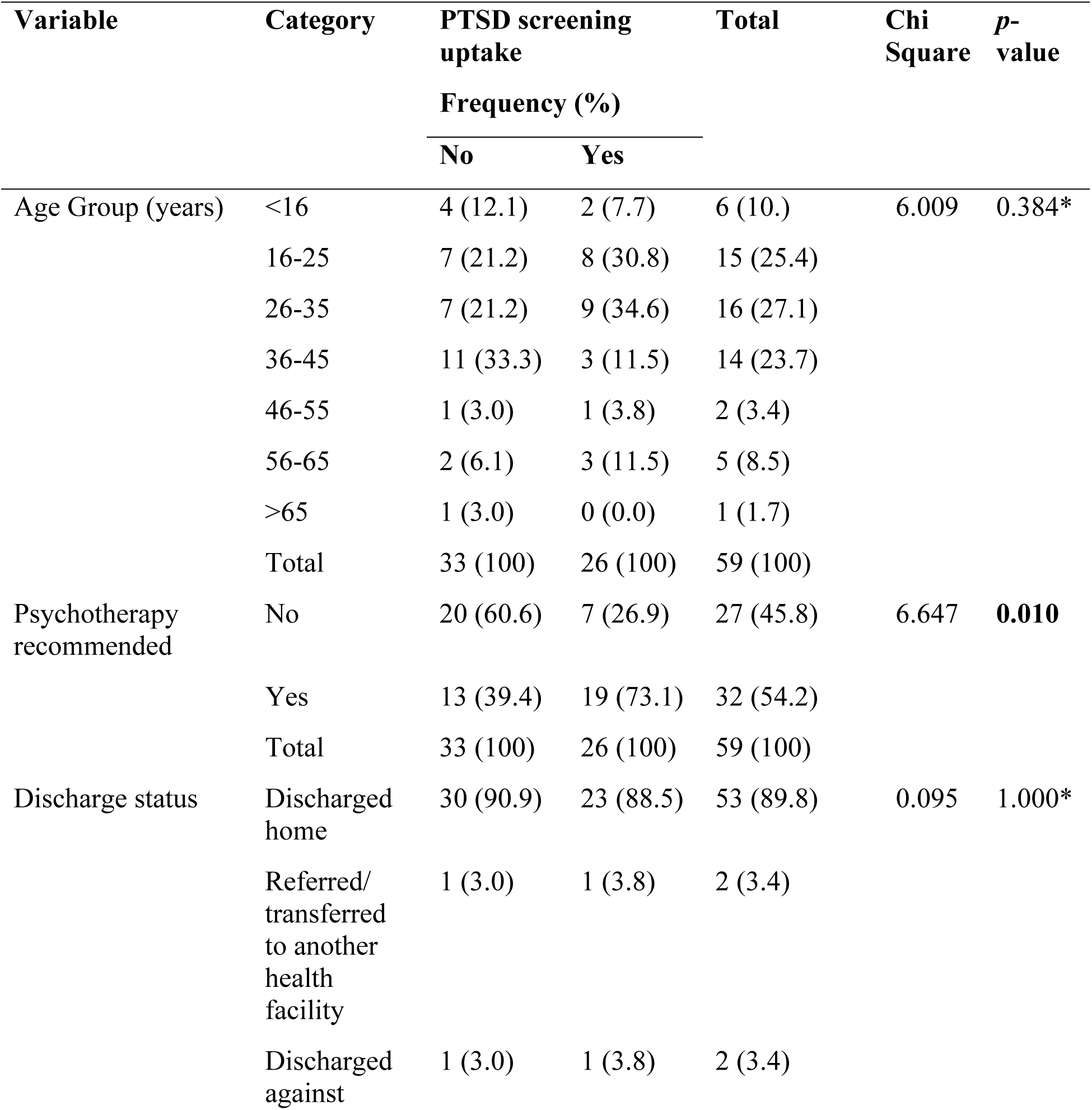
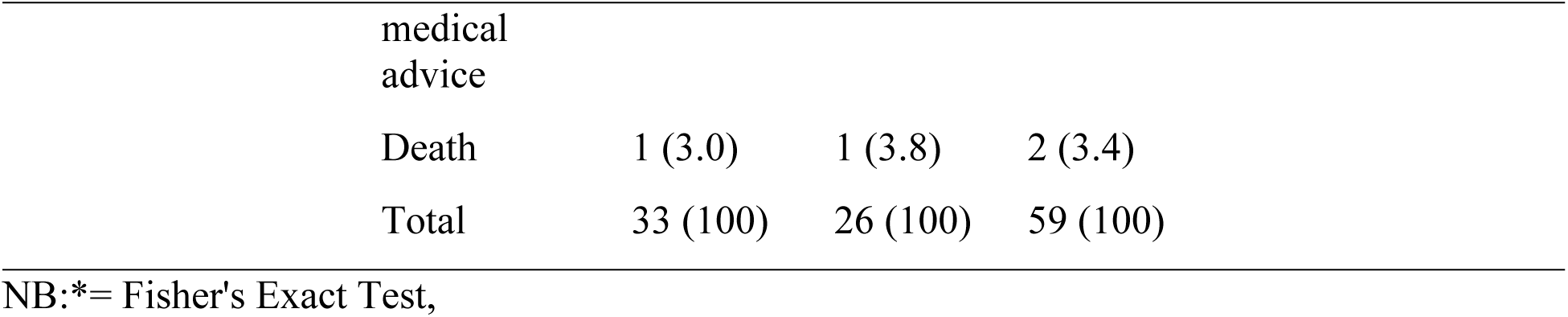
Association between PTSD Screening Uptake after Recommendation, and Demographic, Crash-related and In-Hospital Characteristics.

## Discussion

This study aimed to describe the epidemiological profile of road traffic injury (RTI) patients in hospitals in Fako Division, Cameroon from 2019 to 2023, particularly regarding PTSD screening recommendations and uptake, as a foundation to inform tailored PTSD management strategies. Our study revealed a huge gap in PTSD screening recommendation and PTSD uptake with only 11% among the 534 RTI patients hospitalized being recommended for PTSD screening. PTSD uptake was only 44.1% among the hospitalized RTI patients to whom PTSD screening was recommended. Despite PTSD being a prevalent debilitating mental health condition following RTI, only a few patients were recommended for PTSD screening indicating a significant gap in access to PTSD screening and mental health services for RTI patients in Fako, Division. This highlights the urgent need for improved and holistic care for RTI/trauma patients, taking into account not just patients’ physical needs, but patients’ psychological needs as well. Implementing systematic PTSD screening, and strengthening mental health infrastructure are crucial steps toward providing holistic care and improving outcomes for RTI patients in Cameroon and similar settings globally.

A total of 4218 RTI patient’s records were reviewed. Patients were predominantly males (69.6%). This corroborates previous findings in Cameroon [19, 21], India [29], and Serbia [30]. The high male predominance could be due to the fact that males are more mobile and have a tendency to engage in risk-taking behaviors such as motorbike riding which increases their risk for RTI. The mean age of RTI patients was 30.34 (±13.51) years which is similar to results reported in Yaounde and India where the mean ages reported were; 31years [31] and 32.7 years [29] respectively. However, the mean age in this study is in contrast with the findings of Rajčević and his colleagues in Serbia where the mean age was higher, 41.4 years. This can be explained by the fact that Serbia like other European countries has an ageing population with an average age of 43.9 years in Serbia [30]. Age groups 16-25 and 26-35 were the most involved in RTI. This can be attributed to the fact that youths, especially males are prone to engaging in risky behaviors, such as excessive speed, driving motorcycles for commercial purposes, driving under the influence of alcohol or illicit drugs, and distractions from mobile phone use while driving. Students (19.4%) were among the most involved in RTIs. This contradicts findings from India where students were among the least (9.18%) involved in RTIs [32]. Fako Division has many secondary and higher institutions of learning with students commuting between the towns of the Division, this can be linked to the high proportion of students involved in RTI. More so, majority of students prefer transportation via motorcycles given they are more affordable, increasing their risk of involvement in RTI.

For injury characteristics, we found that road traffic crashes frequently occurred at night (41.7%). This is consistent with findings reported in India of 54% [29] and 61.24% [33] of road crashes occurring at night. A plausible explanation of our finding could be that the population of Fako enjoy nightlife, consequently can indulge in driving under the influence. More so, most people prefer long-distance travels by night via buses such as; the South West-Douala route and the South West-North West route. High traffic of heavy vehicles coupled with high speed, darkness/ absence of street lights along the Tiko-Douala road, poor visibility, road without traffic panels, and poorly marked roads, may have contributed to a high incidence of RTI during this time frame. Motorcycles comprised more than half (53.7%) of the type of vehicle involved in RTIs. This result corroborates studies carried out in the littoral and South West regions of Cameroon [21, 22]. This is probably because most of the motorcycle riders have not undergone formal driving lessons for a motorcycle and majority use motorcycles for business purposes in our context. Dissimilar to our findings, other studies reported motorized four-wheeler as being most involved in RTI [34]. Hence safer means for transportation should be considered in Cameroon other than motorcycles. Ambulance was found to transport just 1.6% of patients to the hospital which is similar to 14.6% cases transported by ambulance reported in New Delhi [29] and 3.9% [6] in Cameroon with data from the Cameroon Trauma Registry, between 2022 and March 2023. This highlights the gap in pre-hospital care/ services provided by Health facilities in Fako for emergency cases such as RTI. Lower limbs (50.6%) and upper limbs (38.9%) were the most affected anatomical locations for the traumatic injury. Similar findings were reported by previous studies in Limbe, Cameroon [35] and India [33]. Our findings contrast that of Rajčević and colleagues in, 2024 in Serbia where head and neck injuries were the most affected parts (37.8%) [30].

With respect to hospital management options received. A total of 534 patients were hospitalized. The number of RTIs requiring hospitalization represents a substantial economic burden in Fako-Cameroon, a LMIC. Only 11.0% of hospitalized cases were recommended for PTSD screening. Of this number PTSD screening uptake was just 44.1%. This clearly shows a huge gap in mental health care, specifically PTSD management, among RTI victims. Healthcare workers are likely to recommend PTSD screening for patients who exhibit symptoms of PTSD and/or distress, which clearly shows the psychological impact of RTI on patients and the need for PTSD screening and timely intervention. This percentage however might be lower and might not reflect the actual number of patients in actual need of PTSD screening as the recommendation for PTSD screening for RTI victims by health providers is subjective and not in routine care. Given PTSD is a prevalent psychological disorder following RTIs, screening of victims one month after road crash should be a routine recommendation to ensure holistic patient care for all victims. This is further compounded by just 8.8% of hospitalized patients recommended for psychotherapy. Psychotherapy is a recommended first line treatment for mental health conditions such as PTSD, and lack of these mental health services in primary care after RTI can likely increase patients’ recovery time, predispose them to comorbid conditions of PTSD like cardiovascular diseases, depression, and substance use. This can reduce patients’ quality of life, increase hospital utilization, and overwhelm the health care system. There is therefore need to incorporate routine timely PTSD screening and evidence-based PTSD management strategies like psychotherapy [34-35] for RTI patients in Primary care in Fako Division and similar settings one month after a road traffic crash.

Bivariate analysis revealed significant associations between; means of transportation of the patient to the hospital, pain medication given, sedative given, surgery performed, psychotherapy recommended and transfusion performed with recommendation for PTSD screening. This could be attributed to the fact that in hospital services provided by health care providers such as; giving pain medication, sedative, surgery performed, psychotherapy recommended, are likely provided to RTI patients with severe conditions who are likely to express more distress prompting health providers to recommend screening for PTSD. The significant associations between the administration of pain medication and sedatives with PTSD screening recommendations could indicate that clinicians recognize that patients requiring higher levels of analgesia and sedation are experiencing significant distress, which may be a trigger for considering psychological evaluation. The significant associations between undergoing surgery or receiving transfusions, and PTSD screening recommendations likely reflect the severity of the physical trauma experienced by these patients. These interventions are typically reserved for more critical cases, where the risk of psychological sequelae like PTSD is also higher. This finding underscores the importance of integrating mental health assessments into the care pathway for patients with severe traumatic injuries. However, this is subjective on the healthcare provider and the level of resilience of the RTI victims and may not reflect the actual number of RTI victims in need of PTSD screening and timely management.

Bivariate analysis for factors associated with PTSD screening uptake revealed a statistically significant association between PTSD screening uptake and recommendation for psychotherapy. The association between recommendation for psychotherapy and PTSD screening uptake highlights the critical role of clinician endorsement in promoting patient engagement with mental health services. Patients are more likely to pursue screening when a healthcare provider explicitly recommends psychotherapy, suggesting that the provider’s recommendation instills confidence and reduces stigma associated with mental health care. Even with a recommendation, actual access to affordable and timely mental health services in Fako Division may be a limiting factor in screening uptake. This underscores the need for training healthcare providers to confidently and routinely recommend screening for PTSD and psychotherapy when appropriate. These findings can be exploited to inform tailored PTSD management strategies. To improve patients, outcomes, it is essential that recommendations for PTSD screening and subsequently psychotherapy be included in routine RTI patients’ care which will increase the likelihood for PTSD screening uptake and early diagnosis as well as treatment. Given the significant association between PTSD screening uptake and recommendation for psychotherapy, patients who go for PTSD screening are likely to be well managed as they will be recommended for psychotherapy which is an evidence based first line treatment for PTSD. Therefore, timely referral for PTSD screening and uptake, timely diagnosis, and recommendation for management by psychotherapy will be ensured leading to improved patients’ outcomes and overall well-being.

Understanding the epidemiological profile of RTI survivors is essential for monitoring and controlling specific factors associated with PTSD screening and uptake. Analyzing hospital data on RTI enables the prioritization of preventive measures and the enhancement of emergency response protocols. These measures may include; the enhancement of emergency medical services such as awareness raising and capacity building of healthcare workers on PTSD management and implementing routine screening for prevalent mental health conditions like PTSD after a road traffic crash. Through collaborative efforts between healthcare authorities, policymakers, and community stakeholders, targeted interventions can be implemented to improve RTI patients’ well-being and quality of life.

## Strengths and limitations

The study’s strengths include a large sample size, multi-hospital data enhancing representativeness, use of real-world clinical data, and a focus on PTSD screening. The limitations include a retrospective design subject to data quality issues, which was mitigated by ensuring that only completed data with variables needed for our study were included.

## Conclusion

Post-traumatic stress disorder screening recommendations and uptake among RTI patients are remarkably low in hospitals in Fako Division. Means of transportation of the patient to the hospital, pain medication given, sedative given, surgery performed, psychotherapy recommended, and transfusion performed were significantly associated with recommendation for PTSD screening Post-traumatic stress disorder screening uptake was significantly associated with recommendation for psychotherapy.

There is a huge need for the development of better trauma care services for RTI patients, especially in-hospital care. An integrated approach geared toward better coordination between units in charge of in-hospital RTI/trauma patient care through better availability of trained human resources (particularly on-the-job capacity building of health care workers) on early identification, screening, and timely management of PTSD among RTI patients is essential. The presence of a good referral system to specialized mental health services (psychologists and psychiatrists) for psychotherapy and pharmacotherapy is crucial to close the gap and improve patients’ outcomes. This study revealed a significant gap in PTSD screening recommendation, uptake, and mental health services for RTI patients in Fako Division, Cameroon, highlighting the need for integrated care and multifaceted interventions. Implementing systematic PTSD screening, timely management, and strengthening mental health infrastructure are crucial steps toward improving outcomes for RTI patients in Cameroon and similar settings globally. Future research should focus on qualitatively exploring the barriers and facilitators to PTSD screening and treatment in this specific context, and on developing targeted interventions to improve access to mental health care, specifically PTSD care, for RTI survivors.

## Abbreviations

RTI: Road traffic injury
PTSD: Post traumatic stress disorder
RTC: Road traffic crash

## Declarations

### Ethics approval

Ethical approval was obtained from the Institutional Review Board of the Faculty of Health Sciences of the University of Buea (No: 2024/2524-04/UB/SG/IRB/FHS) and administrative authorization from the South West Regional Delegation of Public Health.

### Consent for Publication

Not applicable

### Data availability statement

The datasets used and/or analyzed during the current study are available from the corresponding author on reasonable request.

### Acknowledgment

We wish to acknowledge the contributions of the staff and patients at the University of Buea, Buea Regional Hospital, Saint Luke Hospital Buea, and the Limbe Regional Hospital.

### Conflicts of interest

The authors declare that they have no competing interests.

### Funding

Funding was provided by the United States of America Fogarty International Center, National Institutes of Health (NIH) STREaM Cameroon Project, D43TW012186 award.

### Author contributions

CNN: Conceptualization, formal analysis, methodology, writing - original draft; PS: methodology, writing - review & editing; DSN: methodology, writing - review & editing; EAT: formal analysis, review & editing; NT: formal analysis, writing - review & editing; RO: project administration, review & editing; NB: project administration, review & editing; CEU: review & editing; SIM: methodology, review & editing; CJ: Funding acquisition, methodology, review & editing; AC-M: funding acquisition, methodology; EGH-E: methodology, writing - review & editing. All authors read and approved the final manuscript.

## Notes

### Competing Interest Statement

The authors have declared no competing interest.

### Author Declarations

Institutional Review Board of the Faculty of Health Sciences of the University of Buea (No: 2024/2524-04/UB/SG/IRB/FHS)

